# Mental health, health status, and recurrent healthcare utilization in patients with non-obstructive coronary artery disease

**DOI:** 10.1101/2025.11.21.25340776

**Authors:** Paula M.C. Mommersteeg, Nina Kupper, Carmen Siebers, Jos Widdershoven, Paul Lodder

## Abstract

**Background:** Recurrent cardiac healthcare utilization (HCU) is common in patients with non-obstructive coronary artery disease (NOCAD), yet the long-term psychosocial and health-status determinants of care use remain insufficiently understood. Identifying these factors is essential for improving patient-centered management and reducing unnecessary diagnostic testing.

**Methods:** In this 10-year prospective cohort study, 546 patients (52% women) with angiographically or CT-confirmed NOCAD were followed for emergency department (ED) visits for cardiac symptoms and repeat diagnostic cardiac tests. Baseline assessments included measures of mental health (anxiety, depressive symptoms, Type D personality) and health status (fatigue, generic and disease-specific quality of life). Negative binomial regression models examined associations between baseline psychosocial factors and total HCU, adjusting sequentially for age, sex, education, obesity, diabetes, and major adverse cardiovascular events (MACE). Sex-stratified and MACE-stratified exploratory analyses were performed.

**Results:** Half of the cohort experienced at least one cardiac-related healthcare contact during follow-up. In age- and sex-adjusted models, higher baseline anxiety and fatigue, poorer physical health, greater angina frequency, and lower quality of life were associated with higher HCU. After full adjustment, lower baseline quality of life, diabetes, and subsequent MACE remained independent predictors of greater HCU. Sex-stratified analyses showed that more frequent angina and lower quality of life predicted HCU in women but not in men. Most patients with recurrent HCU did not develop MACE, indicating that persistent subjective symptom burden, rather than objective disease progression, drove long-term utilization.

**Conclusions:** In patients with NOCAD, recurrent cardiac HCU is common and is independently predicted by baseline quality of life and cardiometabolic risk. Sex-specific psychosocial patterns contribute to care use, particularly among women. These findings highlight the need to integrate psychosocial assessment, symptom evaluation, and quality-of-life monitoring into routine care to optimize management and reduce recurrent healthcare utilization.

**What Is Known:**

- Recurrent cardiac healthcare utilization (HCU) is common in patients with non-obstructive coronary artery disease (NOCAD), even when objective disease progression is absent.
- Symptom burden and psychosocial distress have been suggested as important drivers of healthcare-seeking behavior in this population.

**What the Study Adds:**

- Lower baseline quality of life independently predicts higher long-term HCU over a 10-year period.
- Sex-specific patterns emerged, with angina frequency and poorer quality of life predicting recurrent HCU in women only.
- Most recurrent care occurred without subsequent major adverse cardiovascular events, underscoring that subjective symptom burden—rather than disease progression—drives long-term utilization.

## Introduction

The majority of patients undergoing coronary angiography (CAG) or computed tomography (CT) for complaints of chest pain have nonobstructive coronary arteries, which may include clean vessels as well as having visible wall irregularities without significant obstructive occlusion (1–3). Such patients may be classified as having ‘No Obstructive Coronary Artery Disease’ (NOCAD), with *persistent* signs and symptoms of chest pain or angina (ANOCA), or, when accompanied by subsequent of myocardial ischemia as ‘Ischemia with No Obstructive CAD’ (INOCA)(4, 5). Treatment for NOCAD focuses on symptom management, such as lifestyle modification and medication (6). However, it has not been that long ago that patients were being discharged as having harmless non-cardiac chest pain (6), while returning frequently, and were thus regarded to ‘utilize a significant part of our health care resources’(7).

Yet, NOCAD remains underdiagnosed and underrecognized, leading to a prolonged and uncertain diagnostic pathway (1, 8, 9). Patients with NOCAD report worse mental health, such as increased anxiety and depressive symptoms, poorer health status, and a higher prevalence of Type D personality, compared to healthy age-matched controls (10, 11), which is more pronounced in women (11, 12). Having persistent cardiac symptoms is associated with recurrent diagnostic tests, and increased health care utilization (HCU) (1, 13). Over time, patients with NOCAD, ANOCA, or INOCA are at increased risk for adverse outcomes, including reduced quality of life, reduced functional capacity and working hours, and major adverse cardiac events (MACE)(3, 13–17). In the present study, we aim to further identify predictors for HCU in patients with NOCAD over a 10 year period. HCU is defined as a combination of the total number of emergency department (ED) visits for cardiac symptoms, and/or total number of undergoing recurrent cardiac diagnostic testing (recurrent test; either a computed tomography (CT) scan, coronary angiography (CAG), a nuclear medicine SPECT-CT, or MRI scan).

The term *frequent attenders* has been coined for people who use disproportionately more care in terms of visits to care facilities compared to a general population, without specifying a particular patient population (18). In their study, Shukla and colleagues have shown that mental health and perceived health were more likely to be associated with frequent care attendance than gender or age (18). Type D personality, a combination of reporting high on negative affectivity and social inhibition, was related to increased HCU in the general population (19). In patients with chronic diseases, a diagnosis of a mental health disorder as well as male sex were related to higher costs of HCU, including hospitalization and emergency department visit rates, length of stay, and hospitalization (20). More specifically for patients with heart disease, psychological distress, particularly depressive symptoms, were associated with greater HCU in patients with coronary artery disease and other chronic diseases (21). Depressive symptoms, anxiety, and poor disease specific quality of life as well as generic health status were related to an increase in health care costs after one year in patients with IHD, adjusted for covariates including gender (22). In a patient group following cardiovascular surgery for over one year, covariate adjusted findings showed that anxiety at baseline was related to more outpatient visits, whereas depressive symptoms were related to more ED visits (23). HCU after a myocardial infarction was related to the personality factor neuroticism, but none of the other personality factors nor depressive symptoms (24). In sum, having worse baseline mental health and health status is hypothesized to be related to recurrent HCU. Although most previous studies adjusted for sex/gender, many did not report sex-stratified findings. Therefore, we explore separately for women and men if worse mental health or health status at baseline is related to increased HCU, hypothesizing associations to be more prevalent in women.

Recurrent HCU is likely to be an indicator of progression of heart disease in patients with NOCAD who experience MACE over time. However, it remains unclear what factors are associated with HCU in patients *without* progression of ischemic heart disease (IHD)(i.e., patients without MACE). In the present study progression of NOCAD is operationalized as the occurrence of MACE, defined as myocardial infarction, percutaneous coronary intervention or coronary artery bypass graft surgery, heart failure, or cardiovascular mortality (16). Moreover, exploring those with eventual progression towards MACE versus no disease progression (no MACE) for baseline mental health and health status could reveal whether those psychological factors are associated with more HCU in those without or with subsequent disease progression.

In sum, the present study investigates the prevalence of recurrent cardiac HCU across a 10-year follow-up period in patients with NOCAD. We aim to examine the predictive associations of baseline mental health and health status with HCU, while adjusting for age and sex, and subsequently for potential covariates, and MACE. We hypothesize that patients with higher scores on baseline depressive symptoms, anxiety, Type D personality, fatigue, and a poor generic and disease specific quality of life will have higher HCU across the follow-up period. We will explore sex-stratified and MACE-stratified associations of mental health and health status for HCU.

## Methods

### Participants and procedure

Data came from the TWeeSteden mIld STenosis (TWIST) study, a single-center prospective cohort on psychosocial risk factors for NOCAD progression (25). Between 2009 and 2013, patients were recruited at ETZ Hospital (Tilburg, NL). Inclusion required a CT calcium score >0 without subsequent CAG referral, or CAG-detected vascular irregularities/mild stenosis; exclusions were normal coronaries, obstructive IHD, prior obstructive events, or insufficient Dutch proficiency. Of 6490 screened, 883 were eligible and invited; 546 (61%) consented and were followed. Questionnaires were completed at baseline, 12, and 24 months; hospital records were reviewed at baseline and ∼10 years. The study was approved by the METC Brabant (NL22258.008.08, amendment ETZ: L1160.2020/LP.2008.227) and conducted in accordance with the Declaration of Helsinki

### Recurrent cardiac health care utilization [HCU]

Recurrent cardiac HCU was retrieved from hospital records (2021/2022)(16, 26). Data included ED visits for cardiac reasons (event and date) and recurrent diagnostic tests (CT, CAG, SPECT, or MRI) since baseline. Total HCU was calculated as the sum of ED visits and retests; events in close succession (e.g., ED visit followed by CAG) were counted separately. Patients without events were censored at the final screening date (Feb 3, 2022); those lost to follow-up were censored 12 months after last contact (16). Time was described as average time between events (or censoring) and maximal study time (baseline CAG/CT to final/censored date).

### Mental health and health status

Validated questionnaires were used to assess mental health and health status. Anxiety and depressive symptoms were measured with the Hospital Anxiety and Depression Scale (HADS; 14 items, subscales 0–21 for anxiety [HADS-A] and depression [HADS-D])(27). Type D personality was assessed with the DS14 (two 7-item subscales: social inhibition [SI] and negative affectivity [NA], range 0–28)(28, 29). Type D was modeled using standardized SI and NA scores plus their interaction (29).

Health status was assessed with the Fatigue Assessment Scale (FAS; 10 items, higher scores = more fatigue) (30), the 12-item Short Form (SF-12; physical [PCS] and mental [MCS] summary scores, higher = better health) (31), and a modified 19-item Seattle Angina Questionnaire (mSAQ) adapted for broader cardiac symptoms (25, 32). The mSAQ measured physical limitation, angina stability, angina frequency, treatment satisfaction, and quality of life(/disease perception), with higher scores indicating better health status (33).

### Covariates and descriptives

At baseline, patients completed a questionnaire on sociodemographics (sex, age, partner status, education, employment status) and lifestyle factors (BMI, obesity, smoking, physical activity, alcohol use). Cardiac risk factors and disease severity were obtained from hospital records: diabetes, hypertension, dyslipidemia, diagnostic exam at inclusion (CAG versus CT scan), visible wall irregularities in three vessels (LAD, CX, RCA), chest pain in the past month, and classification of patients as ANOCA (chest pain symptoms), INOCA (chest pain and detected ischemia), or other (familial/high cardiovascular risk)(34).

### Progression of IHD; Major Adverse Cardiac Events (MACE)

Because recurrent HCU may indicate IHD progression, patients were classified by presence or absence of MACE, used as a proxy for disease progression. MACE, derived from hospital records, included MI, PCI, CABG, heart failure, and cardiovascular mortality (16). Events were recoded into progression (MACE) versus no progression (No MACE). Noncardiac mortality was treated as a censored event and could occur in any subgroup.

### Statistical analyses

For descriptive purposes, three HCU groups were defined: patients who had no HCU since baseline, those with a single HCU (either an ED visit or retest), or all patients with ≥2 HCU events (multiple HCU). The prevalence of events per HCU group was described as the occurrence of recurrent ED visits, retesting, total HCU, MACE (including cardiac and noncardiac mortality), and cardiac and noncardiac mortality. Time since baseline CAG/CT scan, maximum study time, and average time to any event or censoring were also described. Group differences in sociodemographic variables, lifestyle, risk factors, disease severity, mental health, and health status were assessed using Chi-square tests and One-way ANOVA with post-hoc analyses. For variables with ≥2 categories (e.g., alcohol use, NOCAD subgroups), linear-by-linear associations were reported.

To identify covariate predictors of HCU regularized regression was performed in R (*glmnet*). A 10-fold cross-validation determined the optimal alpha (ridge, LASSO, or elastic net) regression. Missing data were imputed using *mice* with predictive mean matching in 10 datasets. Covariates consistently associated with HCU across imputations were included in the final prediction models.

The main outcome was total HCU (count). Due to excess zeros and overdispersion, negative binomial regression with log link was used instead of Poisson regression (35). Mental health and health status variables were tested in separate models, except for Type D traits and their interaction, which were examined together, adjusted for age and sex. Additional covariates were added based on predictors identified through regularized regression, and subsequently for IHD progression dichotomized as no MACE vs. any MACE. Analyses used pooled estimates from five imputed datasets, with factors coded in descending order (reference = 0). Maximum iterations were set to 50.

Explorative analyses were conducted by stratifying the prediction model by sex and stratified for MACE (no versus any) for each mental health and health status factor separately, adjusted for all other covariates. Unless noted otherwise, analyses were run in SPSS 29 (Armonk, NY: IBM Corp). Significance was set at p ≤ .05.

## Results

Follow-up data were available for 546 patients with NOCAD (52% women, mean age 61.4±9.4 years), of whom 44% were classified as INOCA and 38% as ANOCA. Most were included based on CAG screening (70%) versus 30% via CT-scan, and 16% had non-obstructive plaque in three coronary vessels (versus two or less vessels).

### Prevalence of recurrent HCU

Over a median follow-up of 10 years, half of the patients with NOCAD remained event-free (n=273), while 24% (n=130) had a single HCU and 26% (n=143) had multiple HCU. Among the latter, 59% had two events, 22% three, 13% four to six, and 6% seven to sixteen HCU. Descriptive information on the events per HCU group is visualized in Figure 1, and reported in supplemental Table S1, stratified by no HCU, single, and multiple HCU. Overall, 30% had ≥1 ED visit and 36% ≥1 retest. Mean time between events was 5.0 years (SD=1.0) for single HCU and 2.8 years (SD=1.0) for multiple HCU. MACE occurred in 2% of patients without HCU, 12% with single HCU, and 32% with multiple HCU. Cardiovascular mortality occurred in five patients (2%) without prior HCU. Mortality rates (cardiac and non-cardiac) did not differ significantly between HCU groups.

**Figure 1.**
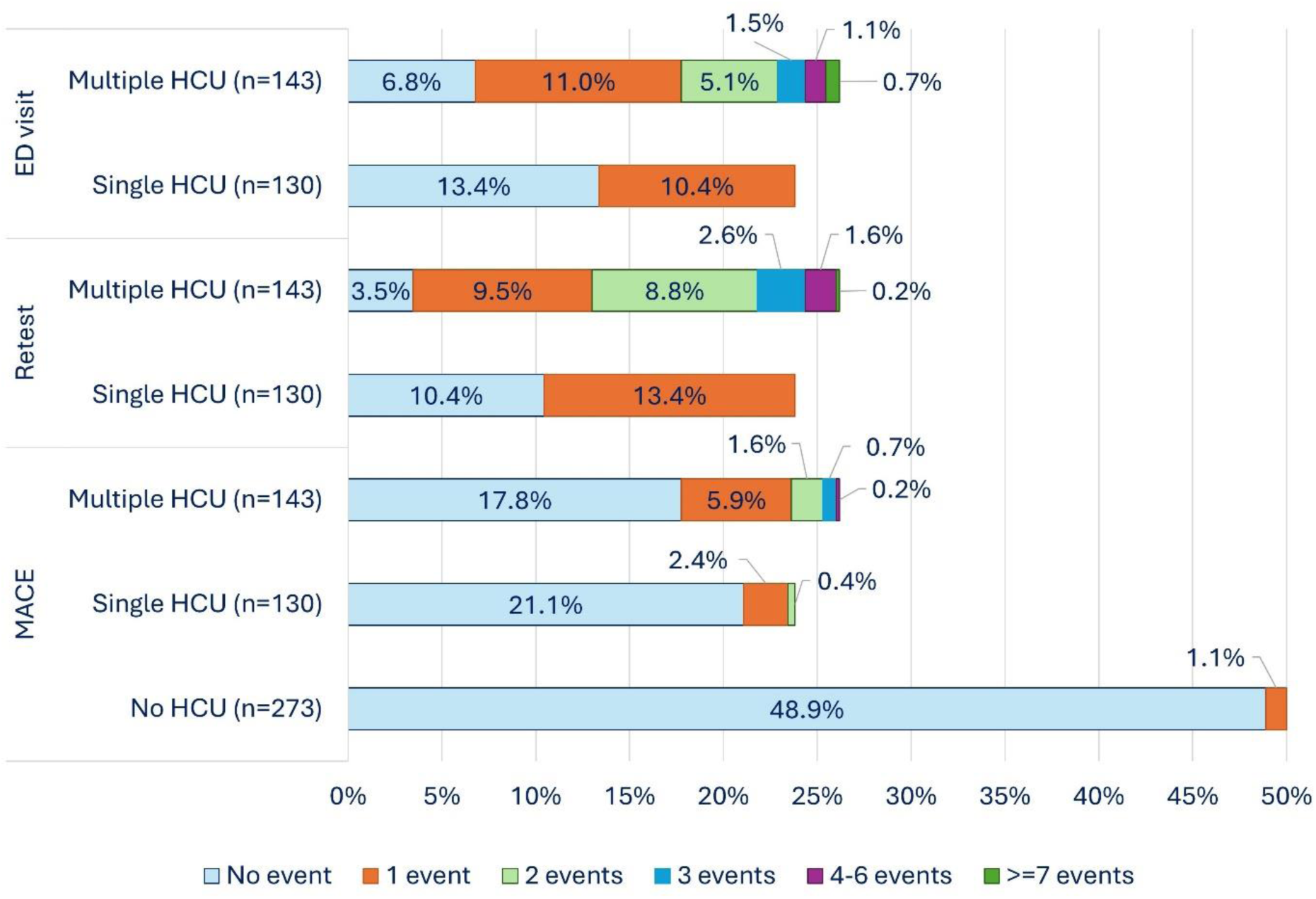
Number of events stratified for ED visit, Retest, and MACE per subgroup of health care utilization.

### Descriptives stratified by HCU groups and covariate selection

No significant sex or age differences were found across HCU groups (Table 1). Patients with multiple HCU had higher anxiety, fatigue, poorer physical health, and lower quality of life. Moreover, the multiple HCU group had less college education, were more often obese, reported more chest pain, and were more often classified as INOCA.

**Table 1.**
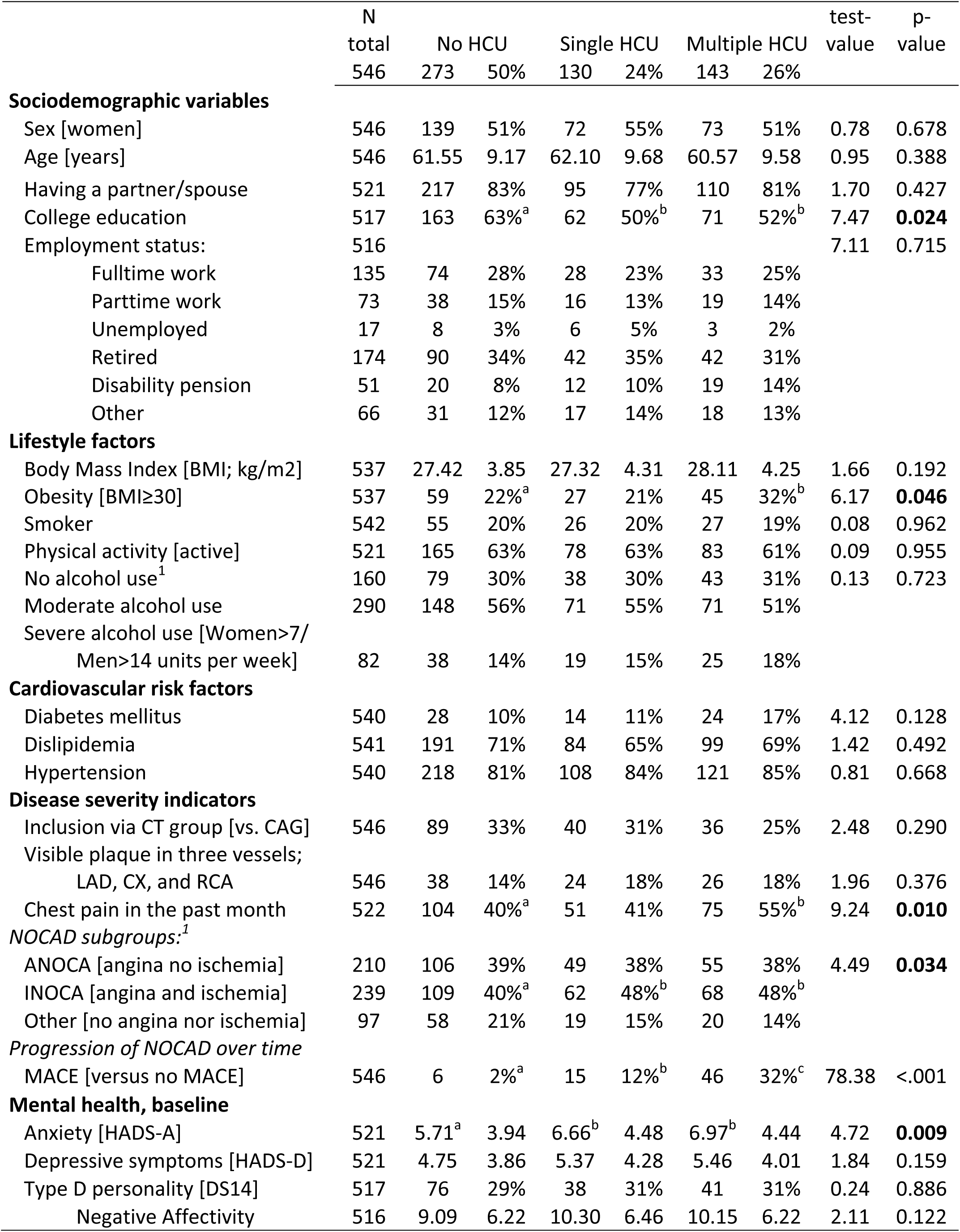
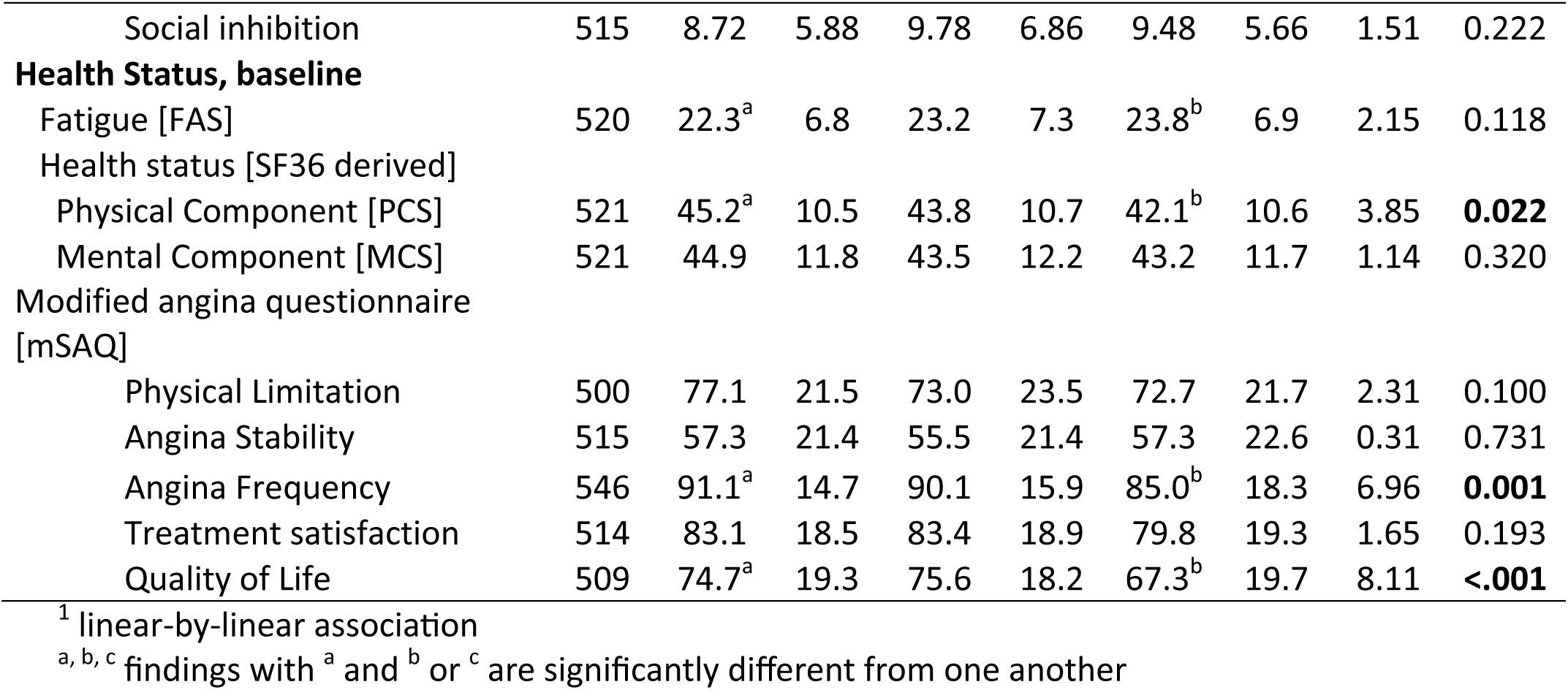
Descriptive variables stratified by HCU groups.

|  | N |  |  |  |  |  |  | test- | p- |
| --- | --- | --- | --- | --- | --- | --- | --- | --- | --- |
|  | total | No HCU |  | Single HCU |  | Multiple HCU |  | value | value |
|  | 546 | 273 | 50% | 130 | 24% | 143 | 26% |  |  |
| **Sociodemographic variables** |  |  |  |  |  |  |  |  |  |
| Sex [women] | 546 | 139 | 51% | 72 | 55% | 73 | 51% | 0.78 | 0.678 |
| Age [years] | 546 | 61.55 | 9.17 | 62.10 | 9.68 | 60.57 | 9.58 | 0.95 | 0.388 |
| Having a partner/spouse | 521 | 217 | 83% | 95 | 77% | 110 | 81% | 1.70 | 0.427 |
| College education | 517 | 163 | 63%<sup>a</sup> | 62 | 50%<sup>b</sup> | 71 | 52%<sup>b</sup> | 7.47 | **0.024** |
| Employment status: | 516 |  |  |  |  |  |  | 7.11 | 0.715 |
| Fulltime work | 135 | 74 | 28% | 28 | 23% | 33 | 25% |  |  |
| Parttime work | 73 | 38 | 15% | 16 | 13% | 19 | 14% |  |  |
| Unemployed | 17 | 8 | 3% | 6 | 5% | 3 | 2% |  |  |
| Retired | 174 | 90 | 34% | 42 | 35% | 42 | 31% |  |  |
| Disability pension | 51 | 20 | 8% | 12 | 10% | 19 | 14% |  |  |
| Other | 66 | 31 | 12% | 17 | 14% | 18 | 13% |  |  |
| **Lifestyle factors** |  |  |  |  |  |  |  |  |  |
| Body Mass Index [BMI; kg/m<sup>2</sup>] | 537 | 27.42 | 3.85 | 27.32 | 4.31 | 28.11 | 4.25 | 1.66 | 0.192 |
| Obesity [BMI≥30] | 537 | 59 | 22%<sup>a</sup> | 27 | 21% | 45 | 32%<sup>b</sup> | 6.17 | **0.046** |
| Smoker | 542 | 55 | 20% | 26 | 20% | 27 | 19% | 0.08 | 0.962 |
| Physical activity [active] | 521 | 165 | 63% | 78 | 63% | 83 | 61% | 0.09 | 0.955 |
| No alcohol use<sup>1</sup> | 160 | 79 | 30% | 38 | 30% | 43 | 31% | 0.13 | 0.723 |
| Moderate alcohol use | 290 | 148 | 56% | 71 | 55% | 71 | 51% |  |  |
| Severe alcohol use [Women>7/<br>Men>14 units per week] | 82 | 38 | 14% | 19 | 15% | 25 | 18% |  |  |
| **Cardiovascular risk factors** |  |  |  |  |  |  |  |  |  |
| Diabetes mellitus | 540 | 28 | 10% | 14 | 11% | 24 | 17% | 4.12 | 0.128 |
| Dislipidemia | 541 | 191 | 71% | 84 | 65% | 99 | 69% | 1.42 | 0.492 |
| Hypertension | 540 | 218 | 81% | 108 | 84% | 121 | 85% | 0.81 | 0.668 |
| **Disease severity indicators** |  |  |  |  |  |  |  |  |  |
| Inclusion via CT group [vs. CAG] | 546 | 89 | 33% | 40 | 31% | 36 | 25% | 2.48 | 0.290 |
| Visible plaque in three vessels;<br>LAD, CX, and RCA | 546 | 38 | 14% | 24 | 18% | 26 | 18% | 1.96 | 0.376 |
| Chest pain in the past month | 522 | 104 | 40%<sup>a</sup> | 51 | 41% | 75 | 55%<sup>b</sup> | 9.24 | **0.010** |
| *NOCAD subgroups:<sup>1</sup>* |  |  |  |  |  |  |  |  |  |
| ANOCA [angina no ischemia] | 210 | 106 | 39% | 49 | 38% | 55 | 38% | 4.49 | **0.034** |
| INOCA [angina and ischemia] | 239 | 109 | 40%<sup>a</sup> | 62 | 48%<sup>b</sup> | 68 | 48%<sup>b</sup> |  |  |
| Other [no angina nor ischemia] | 97 | 58 | 21% | 19 | 15% | 20 | 14% |  |  |
| *Progression of NOCAD over time* |  |  |  |  |  |  |  |  |  |
| MACE [versus no MACE] | 546 | 6 | 2%<sup>a</sup> | 15 | 12%<sup>b</sup> | 46 | 32%<sup>c</sup> | 78.38 | <.001 |
| **Mental health, baseline** |  |  |  |  |  |  |  |  |  |
| Anxiety [HADS-A] | 521 | 5.71<sup>a</sup> | 3.94 | 6.66<sup>b</sup> | 4.48 | 6.97<sup>b</sup> | 4.44 | 4.72 | **0.009** |
| Depressive symptoms [HADS-D] | 521 | 4.75 | 3.86 | 5.37 | 4.28 | 5.46 | 4.01 | 1.84 | 0.159 |
| Type D personality [DS14] | 517 | 76 | 29% | 38 | 31% | 41 | 31% | 0.24 | 0.886 |
| Negative Affectivity | 516 | 9.09 | 6.22 | 10.30 | 6.46 | 10.15 | 6.22 | 2.11 | 0.122 |
| Social inhibition | 515 | 8.72 | 5.88 | 9.78 | 6.86 | 9.48 | 5.66 | 1.51 | 0.222 |
| **Health Status, baseline** |  |  |  |  |  |  |  |  |  |
| Fatigue [FAS] | 520 | 22.3<sup>a</sup> | 6.8 | 23.2 | 7.3 | 23.8<sup>b</sup> | 6.9 | 2.15 | 0.118 |
| Health status [SF36 derived] |  |  |  |  |  |  |  |  |  |
| Physical Component [PCS] | 521 | 45.2<sup>a</sup> | 10.5 | 43.8 | 10.7 | 42.1<sup>b</sup> | 10.6 | 3.85 | **0.022** |
| Mental Component [MCS] | 521 | 44.9 | 11.8 | 43.5 | 12.2 | 43.2 | 11.7 | 1.14 | 0.320 |
| Modified angina questionnaire [mSAQ] |  |  |  |  |  |  |  |  |  |
| Physical Limitation | 500 | 77.1 | 21.5 | 73.0 | 23.5 | 72.7 | 21.7 | 2.31 | 0.100 |
| Angina Stability | 515 | 57.3 | 21.4 | 55.5 | 21.4 | 57.3 | 22.6 | 0.31 | 0.731 |
| Angina Frequency | 546 | 91.1<sup>a</sup> | 14.7 | 90.1 | 15.9 | 85.0<sup>b</sup> | 18.3 | 6.96 | **0.001** |
| Treatment satisfaction | 514 | 83.1 | 18.5 | 83.4 | 18.9 | 79.8 | 19.3 | 1.65 | 0.193 |
| Quality of Life | 509 | 74.7<sup>a</sup> | 19.3 | 75.6 | 18.2 | 67.3<sup>b</sup> | 19.7 | 8.11 | **<.001** |

<sup>1</sup> linear-by-linear association
a, b, c findings with <sup>a</sup> and <sup>b</sup> or <sup>c</sup> are significantly different from one another

Regularized regression identified education, obesity, and diabetes as consistent predictors of HCU, while other disease severity indicators and lifestyle/risk factors were not. These three predictors were added as covariates to the negative binomial GLM, along with age, sex, and MACE.

### Number of HCU by mental health and health status

In separate models examining number of HCU, adjusted for age and sex, anxiety, fatigue, physical component, physical limitation, angina frequency, and quality of life were significantly associated with HCU count (Table 2). More anxiety, fatigue, and poorer health status at baseline were associated with increased HCU. No significant association with HCU was observed for depressive symptoms, Type D personality, mental component, angina stability, and treatment satisfaction. Subsequently, each model was adjusted for education, obesity, and diabetes, showing that angina frequency (B= −0.003, 95%CI −0.006 - −0.0001, p=.041), and quality of life (B= −0.004, 95%CI −0.006 - −0.001, p=.007) remained negatively associated with the number of HCU. A greater angina frequency and a lower quality of life at baseline were related to more HCU, as well as covariates having obesity or diabetes. Finally, in the models adjusted for MACE, MACE (B= 0.558, 95%CI 0.403-0.714, p<.001), quality of life (B= −0.003, 95%CI −0.006 - −0.0003, p=.031), and diabetes (B= 0.141, 95%CI 0.005-0.227, p=.042) were significantly associated with the number of HCU, but none of the other mental health or health status variables nor age, sex, college education, or obesity.

**Table 2.**
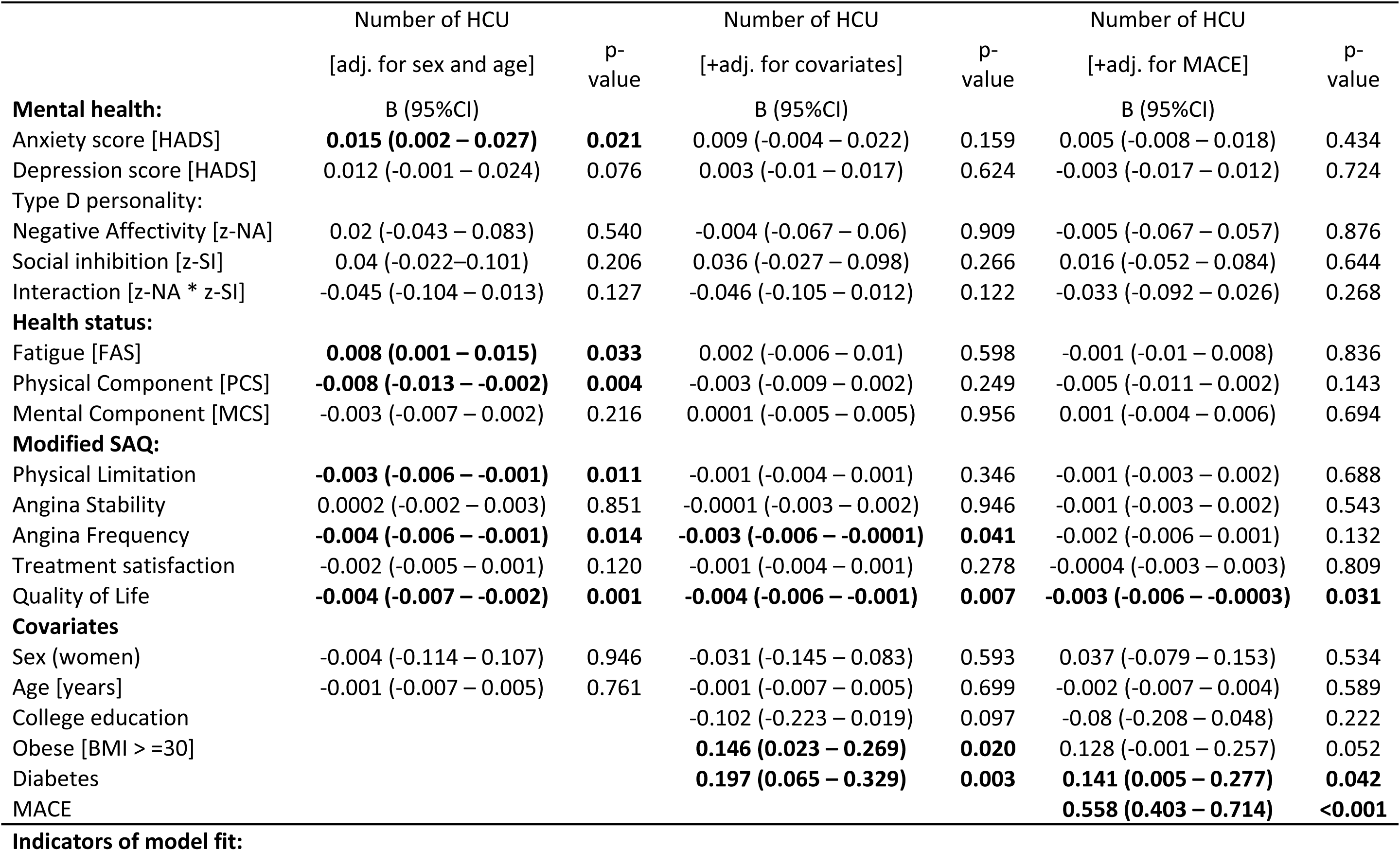
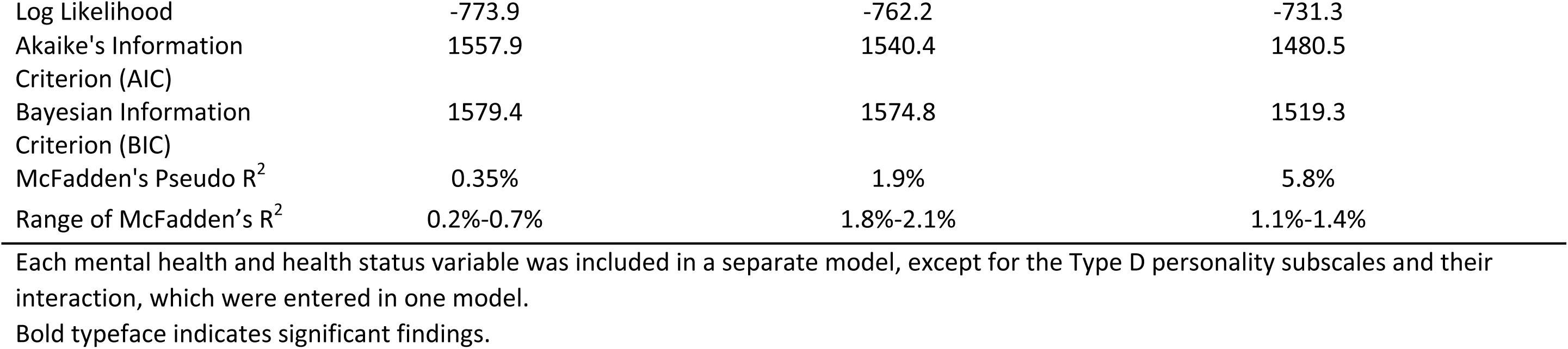
Negative binomial regression of number of HCU events by mental health and health status.

|  | Number of HCU |  | Number of HCU |  | Number of HCU |  |
| --- | --- | --- | --- | --- | --- | --- |
|  | [adj. for sex and age] | p-value | [+adj. for covariates] | p-value | [+adj. for MACE] | p-value |
| **Mental health:** | B (95%CI) |  | B (95%CI) |  | B (95%CI) |  |
| Anxiety score [HADS] | **0.015 (0.002 – 0.027)** | **0.021** | 0.009 (-0.004 – 0.022) | 0.159 | 0.005 (-0.008 – 0.018) | 0.434 |
| Depression score [HADS] | 0.012 (-0.001 – 0.024) | 0.076 | 0.003 (-0.01 – 0.017) | 0.624 | -0.003 (-0.017 – 0.012) | 0.724 |
| Type D personality: |  |  |  |  |  |  |
| Negative Affectivity [z-NA] | 0.02 (-0.043 – 0.083) | 0.540 | -0.004 (-0.067 – 0.06) | 0.909 | -0.005 (-0.067 – 0.057) | 0.876 |
| Social inhibition [z-SI] | 0.04 (-0.022 – 0.101) | 0.206 | 0.036 (-0.027 – 0.098) | 0.266 | 0.016 (-0.052 – 0.084) | 0.644 |
| Interaction [z-NA * z-SI] | -0.045 (-0.104 – 0.013) | 0.127 | -0.046 (-0.105 – 0.012) | 0.122 | -0.033 (-0.092 – 0.026) | 0.268 |
| **Health status:** |  |  |  |  |  |  |
| Fatigue [FAS] | **0.008 (0.001 – 0.015)** | **0.033** | 0.002 (-0.006 – 0.01) | 0.598 | -0.001 (-0.01 – 0.008) | 0.836 |
| Physical Component [PCS] | **-0.008 (-0.013 – -0.002)** | **0.004** | -0.003 (-0.009 – 0.002) | 0.249 | -0.005 (-0.011 – 0.002) | 0.143 |
| Mental Component [MCS] | -0.003 (-0.007 – 0.002) | 0.216 | 0.0001 (-0.005 – 0.005) | 0.956 | 0.001 (-0.004 – 0.006) | 0.694 |
| **Modified SAQ:** |  |  |  |  |  |  |
| Physical Limitation | **-0.003 (-0.006 – -0.001)** | **0.011** | -0.001 (-0.004 – 0.001) | 0.346 | -0.001 (-0.003 – 0.002) | 0.688 |
| Angina Stability | 0.0002 (-0.002 – 0.003) | 0.851 | -0.0001 (-0.003 – 0.002) | 0.946 | -0.001 (-0.003 – 0.002) | 0.543 |
| Angina Frequency | **-0.004 (-0.006 – -0.001)** | **0.014** | **-0.003 (-0.006 – -0.0001)** | **0.041** | -0.002 (-0.006 – 0.001) | 0.132 |
| Treatment satisfaction | -0.002 (-0.005 – 0.001) | 0.120 | -0.001 (-0.004 – 0.001) | 0.278 | -0.0004 (-0.003 – 0.003) | 0.809 |
| Quality of Life | **-0.004 (-0.007 – -0.002)** | **0.001** | **-0.004 (-0.006 – -0.001)** | **0.007** | **-0.003 (-0.006 – -0.0003)** | **0.031** |
| **Covariates** |  |  |  |  |  |  |
| Sex (women) | -0.004 (-0.114 – 0.107) | 0.946 | -0.031 (-0.145 – 0.083) | 0.593 | 0.037 (-0.079 – 0.153) | 0.534 |
| Age [years] | -0.001 (-0.007 – 0.005) | 0.761 | -0.001 (-0.007 – 0.005) | 0.699 | -0.002 (-0.007 – 0.004) | 0.589 |
| College education |  |  | -0.102 (-0.223 – 0.019) | 0.097 | -0.08 (-0.208 – 0.048) | 0.222 |
| Obese [BMI > =30] |  |  | **0.146 (0.023 – 0.269)** | **0.020** | 0.128 (-0.001 – 0.257) | 0.052 |
| Diabetes |  |  | **0.197 (0.065 – 0.329)** | **0.003** | **0.141 (0.005 – 0.277)** | **0.042** |
| MACE |  |  |  |  | **0.558 (0.403 – 0.714)** | **<0.001** |

Mental health and health status HCU in NOCAD
|  |  |  |  |
| --- | --- | --- | --- |
| Log Likelihood | -773.9 | -762.2 | -731.3 |
| Akaike's Information Criterion (AIC) | 1557.9 | 1540.4 | 1480.5 |
| Bayesian Information Criterion (BIC) | 1579.4 | 1574.8 | 1519.3 |
| McFadden's Pseudo R<sup>2</sup> | 0.35% | 1.9% | 5.8% |
| Range of McFadden's R<sup>2</sup> | 0.2%-0.7% | 1.8%-2.1% | 1.1%-1.4% |

Each mental health and health status variable was included in a separate model, except for the Type D personality subscales and their interaction, which were entered in one model.
Bold typeface indicates significant findings.

### Explorative analyses: sex and MACE stratified findings

Explorative sex- and MACE-stratified associations between mental health or health status and HCU are presented in Supplemental Table S2. Among women, angina frequency and quality of life (modified SAQ) were associated with HCU, whereas no significant associations were observed in men. These sex-stratified models were adjusted for age, education, obesity, diabetes, and MACE. In women, both MACE and obesity, and in men, MACE, lower education, and diabetes were consistently associated with HCU (data not shown).

In patients without MACE, higher anxiety, poorer physical health, greater angina frequency, and lower quality of life at baseline were associated with higher HCU. In patients with MACE, higher depressive symptoms, more fatigue, and poorer physical and mental health status were associated with more HCU. These MACE-stratified models were adjusted for sex, age, education, obesity, and diabetes. Among covariates, obesity and diabetes were consistently related to higher HCU, but only in patients with MACE (data not shown).

## Discussion

In this 10-year follow-up study of patients with non-obstructive coronary artery disease (NOCAD), recurrent cardiac healthcare utilization (HCU) was frequent, with half of patients experiencing one or more cardiac-related contacts. Higher baseline anxiety, fatigue, poorer physical health, and lower disease-specific quality of life were linked to greater HCU, although most associations weakened after adjustment. Quality of life at baseline emerged as an independent predictor, together with diabetes and the occurrence of major adverse cardiovascular events over time (MACE). Explorative analyses suggested these associations were more pronounced in women and different in those who did or did not developed MACE, underscoring potential sex differences and the interplay between psychosocial and clinical factors. To contextualize these findings, we first describe the prevalence and patterns of recurrent HCU among patients with NOCAD.

### Prevalence of HCU events in NOCAD

HCU was assessed as a composite outcome of all cardiac emergency department (ED) visits and repeat diagnostic cardiac tests over 10 years, to capture the recurrent and persistent nature of care use in this population. The high prevalence observed aligns with previous reports of persistent chest pain in NOCAD (5, 36).

In a retrospective cohort, Roguin and colleagues reported a lower recurrence (10% repeat CAG; 3% ischemic heart disease progression) with higher rates among men and those with cardiometabolic risk factors (37). In contrast, the present TWIST study prospectively included patients with any visible wall irregularities, representing a group with more pronounced coronary abnormalities. This broader inclusion yielded a higher prevalence (36%) of recurrent testing, not limited to CAG. Our results partly align with earlier studies showing repeat testing was more common among patients with minimal disease, diabetes, older age, and male sex (17, 38, 39). Overall, recurrent cardiac HCU is common in NOCAD and appears driven by a combination of clinical risk factors and psychosocial mechanisms explored below.

### Mental health and health status at baseline predicting HCU

In age- and sex-adjusted analyses, higher anxiety and fatigue, poorer physical health, greater disease-specific limitation, more frequent angina, and lower quality of life were associated with greater HCU. After full adjustment, only lower quality of life remained an independent predictor. These findings align with prior research linking mental health and physical symptom burden to healthcare use in IHD (21, 23))(22). Experiencing depressive symptoms, in patients with obstructive CAD or who received cardiac surgery, was associated with increased ED visits and overall HCU (21, 23). A one-year follow-up of patients with IHD previously showed that anxiety, depression, and poorer cardiac-specific and generic health status were associated with higher healthcare costs (22). This emphasizes the intertwined role of psychological and physical functioning in care-seeking behaviour.

Explorative MACE-stratified analyses suggested that psychosocial correlates of HCU differ by disease progression. While anxiety, angina frequency, and lower quality of life were associated with HCU in patients *without* MACE, depressive symptoms, fatigue, and poorer physical and mental health were relevant in patients with MACE. Thus suggesting that psychosocial drivers of recurrent care may differ between patients with and without objective progression.

Most studies have examined QoL as an outcome variable. Our findings extend this literature by identifying it as a predictor of recurrent care use, independent of disease severity. Poor QoL may therefore represent a key driver of healthcare-seeking in NOCAD, beyond traditional risk factors or mental health symptoms.

Although no overall sex differences were observed, exploratory analyses suggested sex-specific patterns. In women, higher angina frequency and lower QoL predicted greater HCU, while no significant associations were found in men. This may reflect heightened symptom perception or concern about recurrence in the absence of clear disease markers, consistent with evidence linking anxiety to increased care-seeking in women with NOCAD and noncardiac chest pain (40, 41). Recent findings by Liu et al. (2024) support this interpretation. Among women with angina and no obstructive CAD (ANOCA), psychological distress was significantly elevated compared with controls, independent of objective ischemia (10). Notably, women with milder angina reported the highest levels of anxiety, depression, and perceived stress, suggesting that subjective symptom experience—rather than ischemic burden—may drive distress and recurrent care-seeking (10). Taken together, these findings point toward sex-specific psychosocial mechanisms underlying HCU in NOCAD, warranting further study and targeted psychosocial support.

### Disease progression and covariates associated with HCU

Experiencing MACE was associated with greater HCU, as such events are often preceded by ED visits or diagnostic testing. However, a small proportion of patients (2%, n=6) experienced MACE without preceding HCU, including five who died from cardiovascular causes. Conversely, the majority of patients with one or multiple HCU episodes (88% and 68%, respectively) did *not* experience disease progression, highlighting that NOCAD burden extends beyond the pathway toward obstructive IHD. Recurrent HCU thus represents a meaningful dimension of patient burden.

Consistent with prior studies, baseline diabetes independently predicted higher HCU (37, 38). Obesity also showed an initial association, suggesting cardiometabolic factors contribute to symptom persistence and repeated testing. Vatsa et al. (2024) reported that obesity was associated with more frequent and severe chest pain, partly mediated by depressive symptoms (36), supporting this interpretation. Neither sex nor age were significantly related to HCU in our study, aligning with some (17, 37), but not all previous studies (37, 38).

### Limitations

Several limitations should be considered. HCU was defined as cardiac ED visits or repeat diagnostic testing; other encounters, such as annual check-ups or arrhythmia assessments, were not included, potentially underestimating total healthcare utilization. Unlike studies defining hospitalization as admission ≥24 hours, our definition captured acute or diagnostic events possibly related to IHD progression. Each hospital contact was categorized as an ED visit or retest, irrespective of disease progression, which was analyzed separately.

Second, patient inclusion (2008–2013) preceded widespread use of coronary function testing for vasospasm or microvascular dysfunction (42, 43). Though ANOCA/INOCA could not be confirmed based on coronary function test, patients were categorized by presenting features (angina-like symptoms or detected ischemia on functional testing). The INOCA group was significantly associated with HCU compared to the ‘other’ group, but other covariates had a stronger association with HCU.

Finally, residual confounding and sample size constraints may have limited power in sex- and MACE-stratified analyses, despite the study’s 10-year longitudinal design.

### Implications

Patients with signs of IHD but NOCAD remain a diagnostic and therapeutic challenge, contributing substantially to healthcare use. Poor baseline QoL independently predicted recurrent cardiac HCU, emphasizing the importance of addressing symptom burden and psychosocial factors in long-term management.

Recent evidence indicates that invasive coronary function testing can identify microvascular dysfunction or vasospasm, providing both prognostic and cost-effective benefits (44–46). The 2024 European Society of Cardiology Chronic Coronary Syndromes guidelines recommend such testing in patients with refractory angina and suspected ANOCA/INOCA to guide individualized treatment (47). Improving recognition and management of these conditions in both primary and specialist care—through better symptom assessment and early initiation of targeted therapy—may reduce recurrent healthcare use and improve patient well-being, even without invasive testing (8, 9).

## Conclusions

This 10-year follow-up study demonstrates that recurrent cardiac HCU is common among patients with NOCAD and is largely driven by QoL rather than disease progression. These findings highlight the need for patient-centered approaches focusing on symptom management, psychosocial well-being, and early identification of functional coronary disorders to reduce recurrent care and improve long-term QoL.

## Acknowledgement

We are indebted to and inspired by the patients who participated in the TWIST study. Many students and colleagues have collaborated for short or longer time in the TWIST project. Each has been acknowledged or coauthored in a previous research paper. We are grateful for the ETZ science office, ETZ cardiology department, and ETZ IT support.

## Funding

This research received no specific grant from any funding agency in the public, commercial, or not-for-profit sectors

## Disclosure of interest

The authors declare no conflicts of interest related to this work.

## Ethics statement

The study was conducted in accordance with the Declaration of Helsinki. Ethical approval was obtained from the institutional review board of METC Brabant (NL22258.008.08, amendment ETZ: L1160.2020/LP.2008.227), and all participants provided written informed consent prior to inclusion in the study.

## Data availability statement

The data underlying this article are available upon reasonable request to the corresponding author. The data are not publicly available due to privacy and ethical restrictions.

## Supplemental Material

Supplemental Table S1: prevalence of events per HCU group

Supplemental Table S2: Sex and MACE stratified psychosocial factors associated with number of HCU, adjusted for covariates

---

|  | Item No. | Recommendation | Page No. | Relevant text from manuscript |
| --- | --- | --- | --- | --- |
| Title and abstract | 1 | (a) Indicate the study's design with a commonly used term in the title or the abstract | 2 | abstract |
|  |  | (b) Provide in the abstract an informative and balanced summary of what was done and what was found | 2-3 | abstract |
| **Introduction** |  |  |  |  |
| Background/rationale | 2 | Explain the scientific background and rationale for the investigation being reported | 4-6 | Introduction |
| Objectives | 3 | State specific objectives, including any prespecified hypotheses | 6 | Introduction |
| **Methods** |  |  |  |  |
| Study design | 4 | Present key elements of study design early in the paper | 7 | Participants and procedure |
| Setting | 5 | Describe the setting, locations, and relevant dates, including periods of recruitment, exposure, follow-up, and data collection | 7 | Participants and procedure |
| Participants | 6 | (a) *Cohort study*—Give the eligibility criteria, and the sources and methods of selection of participants. Describe methods of follow-up<br>*Case-control study*—Give the eligibility criteria, and the sources and methods of case ascertainment and control selection. Give the rationale for the choice of cases and controls<br>*Cross-sectional study*—Give the eligibility criteria, and the sources and methods of selection of participants | 7 | Participants and procedure |
|  |  | (b) *Cohort study*—For matched studies, give matching criteria and number of exposed and unexposed<br>*Case-control study*—For matched studies, give matching criteria and the number of controls per case | n.a. |  |
| Variables | 7 | Clearly define all outcomes, exposures, predictors, potential confounders, and effect modifiers. Give diagnostic criteria, if applicable | 7-8 |  |
| Data sources/<br>measurement | 8* | For each variable of interest, give sources of data and details of methods of assessment (measurement). Describe comparability of assessment methods if there is more than one group | 7-8 |  |
| Bias | 9 | Describe any efforts to address potential sources of bias | 9-10 | Statistical analyses |
| Study size | 10 | Explain how the study size was arrived at | 7 | Participants and procedure |
| Continued on next page<br>Quantitative variables | 11 | Explain how quantitative variables were handled in the analyses. If applicable, describe which groupings were chosen and why | 7-10 | methods |
| Statistical methods | 12 | (a) Describe all statistical methods, including those used to control for confounding | 9-10 | Statistical analyses |
|  |  | (b) Describe any methods used to examine subgroups and interactions | 9-10 | Statistical analyses |
|  |  | (c) Explain how missing data were addressed | 9-10 | Statistical analyses |
|  |  | (d) *Cohort study*—If applicable, explain how loss to follow-up was addressed | 9-10 | Statistical analyses |
|  |  | *Case-control study*—If applicable, explain how matching of cases and controls was addressed |  |  |
|  |  | *Cross-sectional study*—If applicable, describe analytical methods taking account of sampling strategy |  |  |
|  |  | (e) Describe any sensitivity analyses | 9-10 | Statistical analyses |
| **Results** |  |  |  |  |
| Participants | 13* | (a) Report numbers of individuals at each stage of study—eg numbers potentially eligible, examined for eligibility, confirmed eligible, included in the study, completing follow-up, and analysed | 7, 11 | Participants and procedure; results |
|  |  | (b) Give reasons for non-participation at each stage | 7, 11 | Participants and procedure; results |
|  |  | (c) Consider use of a flow diagram | n.a. |  |
| Descriptive data | 14* | (a) Give characteristics of study participants (eg demographic, clinical, social) and information on exposures and potential confounders | Page 11-13<br>Page 23, 24 | Results; Table 1, also supplemental table S1 |
|  |  | (b) Indicate number of participants with missing data for each variable of interest | Page 11-13<br>Page 23, 24 | Results; Table 1, also supplemental table S1 |
|  |  | (c) *Cohort study*—Summarise follow-up time (eg, average and total amount) |  |  |
| Outcome data | 15* | *Cohort study*—Report numbers of outcome events or summary measures over time | Page 11-13<br>Page 23, 24 | Results; Table 1, also supplemental table S1 |
|  |  | *Case-control study*—Report numbers in each exposure category, or summary measures of exposure |  |  |
|  |  | *Cross-sectional study*—Report numbers of outcome events or summary measures |  |  |
| Main results | 16 | (a) Give unadjusted estimates and, if applicable, confounder-adjusted estimates and their precision (eg, 95% confidence interval). Make clear which confounders were adjusted for and why they were included | Page 11-13<br>Page 23, 24 | Results; Table 1, also supplemental table S1 |
|  |  | (b) Report category boundaries when continuous variables were categorized | Page 11-13 | Results; Table 1, also supplemental |
|  |  |  | Page 23, 24 | table S1 |
|  |  | (c) If relevant, consider translating estimates of relative risk into absolute risk for a meaningful time period | n.a. |  |
| Continued on next page |  |  |  |  |
| Other analyses | 17 | Report other analyses done—eg analyses of subgroups and interactions, and sensitivity analyses | Page 12, 13 | Results and Supplemental Table S2: Sex and MACE stratified psychosocial factors associated with number of HCU, adjusted for covariates |
| **Discussion** |  |  |  |  |
| Key results | 18 | Summarise key results with reference to study objectives | 14 | Discussion |
| Limitations | 19 | Discuss limitations of the study, taking into account sources of potential bias or imprecision. Discuss both direction and magnitude of any potential bias | 17 | Limitations |
| Interpretation | 20 | Give a cautious overall interpretation of results considering objectives, limitations, multiplicity of analyses, results from similar studies, and other relevant evidence | 14-18 | Discussion |
| Generalisability | 21 | Discuss the generalisability (external validity) of the study results | 17-18 | Implications, conclusion |
| **Other information** |  |  |  |  |
| Funding | 22 | Give the source of funding and the role of the funders for the present study and, if applicable, for the original study on which the present article is based | 19 | Funding |

\*Give information separately for cases and controls in case-control studies and, if applicable, for exposed and unexposed groups in cohort and cross-sectional studies.
**Note:** An Explanation and Elaboration article discusses each checklist item and gives methodological background and published examples of transparent reporting. The STROBE checklist is best used in conjunction with this article (freely available on the Web sites of PLoS Medicine at http://www.plosmedicine.org/, Annals of Internal Medicine at http://www.annals.org/, and Epidemiology at http://www.epidem.com/). Information on the STROBE Initiative is available at www.strobe-statement.org.

## Notes

### Competing Interest Statement

The authors have declared no competing interest.

### Clinical Trial

NCT01788241

